# Equity Readiness Index for Digital Health Organizations

**DOI:** 10.1101/2025.11.14.25340230

**Authors:** Laura Wagon, Marcia Nißen, Tobias Kowatsch

## Abstract

**Objective:** As healthcare becomes increasingly digitalized, new challenges and opportunities arise for advancing health equity. In response, a broad range of frameworks and recommendations have emerged to guide equitable innovation. Yet despite this momentum, real world progress remains limited and many digital health organizations (DHOs) struggle to translate health equity principles into concrete, actionable strategies. One promising avenue for closing this implementation gap lies in the use of organizational readiness assessments that not only diagnose current capacity but also help to define a roadmap for improvement. In response, this study introduces the *Health Equity Readiness Index* and a supporting novel self-assessment tool designed to support DHOs in evaluating and advancing their readiness to act on health equity.

**Methods:** Drawing on established frameworks from health equity, responsible AI, digital innovation, and organizational change, we created a five-dimensional *Health Equity Readiness Index*, encompassing strategy, governance, culture, data, and community collaboration. Each dimension was operationalized into 15 readiness indicators across four levels. A corresponding digital self-assessment tool was developed and refined through user testing and a structured online survey targeting DHOs.

**Results:** The resulting tool provides a structured, low-barrier mechanism for DHOs to assess their current equity capacity and identify priority areas for improvement. Survey results (N = 124) showed broad applicability across diverse organizational contexts, with respondents spanning regions, service types, and roles. Participants rated the tool highly across all constructs (M ≥ 4.39/5). Qualitative feedback highlighted five overarching strengths (clarity, diagnostic value, actionability, useability, awareness) and three areas for future improvement (examples, customization, additional features). Overall, respondents perceived the tool as both relevant and actionable for guiding equity-oriented strategy.

**Conclusion:** This study contributes a novel framework and diagnostic tool for advancing organizational readiness for health equity in digital health, supporting DHOs in moving from intention to implementation. It also offers potential utility for funders, regulators, and health systems seeking to institutionalize equity through incentives or benchmarking.

## INTRODUCTION

As healthcare becomes increasingly digitalized, new challenges and opportunities arise for advancing health equity [1]. On the one hand, digital health technologies (DHTs) have the potential to transform healthcare by increasing access to care, enabling personalized care, and bridging geographic and socioeconomic gaps in service delivery [2–5]. On the other hand, without intentional safeguards, DHTs risk exacerbating existing inequities by deepening the digital divide for populations with limited access to technology[6], lower digital (health) literacy[7, 8], or lower socioeconomic status[8–10]. These risks are further magnified by the integration of artificial intelligence (AI) due to inherent biases [11]. Bias in AI can arise from three main sources: the use of historical data that reflects and perpetuates systemic inequities [11–15], algorithmic design choices that produce inequitable outcomes for marginalized populations [16–18], and the ways clinicians and patients interact with AI tools [16]. These challenges emphasize the importance of developing and deploying AI systems that are fair, inclusive, and transparent, also referred to as responsible AI [19]. As (AI-driven) DHTs become increasingly embedded in health systems worldwide, addressing their equity implications is no longer optional but essential [4, 20–24].

In response, a growing number of works have emerged on how to advance health equity in digital health [1]. Several frameworks have introduced strategies to guide the design and implementation of equitable DHIs, e.g., through human-centered design [25], participatory research approaches [26], or equity by design principles [27]. Other works have examined how DHIs can be developed and deployed to promote equitable access and outcomes across diverse populations, including those marginalized by race, ethnicity, or socioeconomic status [28–31]. Conceptual advances have sought to modernize established health disparity frameworks for the digital age, for instance, by broadening traditional health equity models to account for digital determinants of health [22] or by incorporating digital access into the broader landscape of social determinants of health [21]. Overall, these efforts provide a strong foundation for equity in digital health.

Despite the growing recognition of health equity as a priority, real-world progress has been limited. In the United States, for example, little measurable improvement has been made in reducing disparities over the past 25 years [32]. Beyond ethical implications, this contributes to billions of dollars in preventable annual healthcare costs, driven by avoidable hospitalizations, poor chronic disease management, and suboptimal outcomes [33] and the cost of inaction on health equity continues to rise globally [34]. Simultaneously, investments in health, particularly those aimed directly at reducing inequities, have long demonstrated high financial returns, driven by gains in human capital and reductions on downstream expenditures [34]. As healthcare systems globally transition towards value-based care (VBC) [35, 36], the business case for addressing equity further strengthens. VBC ties payment to health outcomes [36], meaning organizations that fail to reach and treat marginalized populations effectively might face financial repercussions. In Germany, for example, all prescription digital therapeutics will be obliged to add performance-related components to their reimbursement agreements starting January 2026 [37]. As such, health equity is no longer just an ethical aspiration but is becoming a strategic necessity [38, 39]. However, translating health equity recommendations into actionable strategies remains a persistent challenge for digital health organizations (DHOs). Common barriers include limited internal expertise, data challenges such as collecting and analyzing equity-relevant data, competing priorities, and a lack of practical tools to guide prioritization and implementation within resource-constrained environments [27].

One promising avenue lies in the use of organizational readiness assessments and, as a result, in defining an implementation roadmap to address identified shortcomings. These tools provide structured pathways for diagnosing current capabilities, prioritizing improvements, and building sustained capacity for change [40]. While such approaches are well-established in fields like digital transformation [41], cybersecurity [42], and (responsible) AI [11, 43, 44], they remain underdeveloped in the context of health equity. Furthermore, existing academic models have been criticized for lacking actionable guidance and supportive tools, such as software or online toolkits, that facilitate their real-world implementation [45].

In response, this research introduces the ***Health Equity Readiness Index***, a novel self-assessment approach for DHOs. The study answers the following research question: *How can digital health organizations assess and strengthen their readiness to implement health equity recommendations?*

Grounded in interdisciplinary maturity models and adapted through a health equity lens, the readiness index enables organizations to evaluate their internal capacity to advance equity and identify priority areas for action. In doing so, it addresses a critical gap between knowledge and implementation, offering a practical tool to support sustained, equity-oriented organizational transformation.

## METHODS

The development of the readiness index was inspired by the maturity model development framework by de Bruin and Rosemann [46]. First, we constructed a health equity readiness matrix, drawing on existing frameworks on health equity, responsible AI, and digital innovation, to define the capabilities required to embed equity in DHOs. We then derived and deployed a self-assessment tool that allows organizations to evaluate their current equity readiness, including a curated set of recommendations to improve their readiness index on various dimensions of the readiness matrix.

### Health Equity Readiness Matrix

The foundation of any readiness model lies in its readiness matrix, composed of defined dimensions and progression levels [47]. To define the health equity readiness matrix, we adopted a derivative approach, adapting constructs from established readiness and maturity frameworks to the context of health equity in digital health [47].

### Readiness dimensions

Readiness dimensions form the conceptual core of any readiness model, as they define the key organizational capabilities that distinguish one level of readiness from another [45]. To identify and define the readiness dimensions for our model, we conducted a targeted synthesis of the following four foundational works:

**Marcus et al. [48]** conducted a scoping review of organizational health equity capacity assessments (OCAs) used by public health organizations. OCAs are increasingly recognized as valuable tools for understanding and enhancing an organization’s readiness and capacity to advance health equity [49]. Across the included studies, all OCAs consistently assessed the following internal organizational capabilities: *institutional leadership and governance, budget alignment and resource allocation, commitment and shared visions, internal structures, use of data, staff training, support, and/or staff diversity*. A subset of OCAs also examined external-facing capacities, including the *strength of collaboration with community partners*.

**Doherty et al. [50]** developed a framework for organizational change to advance health equity by adapting Kotter’s eight-step model [51] for leading change and integrating insights from a prior scoping review. This framework was further refined through a qualitative interview study involving 19 experts in 2020. The study identified six core components essential for embedding health equity within healthcare systems: *committed and engaged leadership, integrated organizational structure, commitment to quality improvement and patient safety, ongoing training and education, effective data collection and analytics*, and *stakeholder communication, engagement, and collaboration*

**Yams et al. [44]** developed a multi-dimensional AI maturity model, identifying essential organizational conditions and capabilities required to integrate AI into existing systems. The model comprises six interrelated and interdependent dimensions: strategy, ecosystems, mindsets, organizational structures and processes, data, and technology. Although it does not explicitly address health equity, its relevance lies in the growing role of AI-driven innovation in healthcare [52, 53]. As such, AI innovation maturity can be viewed as adjacent and transferable.

**Willems [54]** developed a responsible AI maturity model tailored to public sector organizations. The model is grounded in a systematic review of 22 studies and refined through a three-round Delphi study. They outline six maturity dimensions: *strategy, culture and competences, organization and management, governance and processes, data and information*, and *technology and tooling*. Responsible AI outlines organizational practices designed to mitigate AI-related risks, including bias, discrimination, and privacy breaches, all of which can carry significant societal implications and may exacerbate existing inequities if left unaddressed [19]. While the model does not explicitly target health equity, the emphasis on fairness, transparency, and accountability in AI aligns closely with equity goals, particularly in health systems increasingly reliant on data-driven technologies.

These frameworks were selected based on their relevance to organizational capacity for health equity, digital innovation, and responsible AI, collectively covering both the organizational and technological dimensions of DHOs. We conducted a comparative analysis across the four foundational frameworks. Each source was reviewed for core constructs (e.g., strategy, culture, data use) and mapped to emerging thematic categories. Converging concepts were abstracted and grouped iteratively, resulting in five high-level readiness dimensions: (1) strategy & organizational commitment, (2) governance & processes, (3) culture & competencies, (4) data & technology, and (5) community collaboration.

### Readiness levels

Readiness levels describe the developmental progression of an organization’s capacity to implement equity-oriented practices. Consistent with best practices in maturity model design [47], we adopted a four-level structure for simplicity and relevance, starting with regulatory requirements and industry best practices, followed by increasing integration of voluntary best practices. Since there is no standard naming convention for readiness models [45], we opted for a naming convention that mirrors the gradual evolution from non-recognition to full institutionalization of equity practices: (1) Unrecognized Need, (2) Emerging Awareness, (3) Developing Capacity, and (4) Established Practices.

### Matrix population

Finally, to derive the full health equity readiness matrix, level descriptors were adapted from [44] and [54] and recontextualized through an equity lens. For instance, “use of data” was adapted to emphasize disaggregation, intersectionality, and equity metrics. Sample descriptors for the dimension *strategy & organizational commitment* are provided in Supplementary Material SM.1.

### Equity Readiness Self-Assessment Tool

#### Tool design

To operationalize the health equity readiness matrix into a practical diagnostic tool, we developed a self-assessment designed to evaluate an organization’s current level of readiness to implement equity-focused strategies. The tool translates the readiness matrix into a structured set of 15 items (see SM.2). Each item presents a question targeting a specific organizational capability and was directly derived from the five readiness dimensions to reflect the multi-level structure of the underlying matrix. Most dimensions are represented by three sub-dimensions and thus three corresponding items, except for Data & Technology (four items) and Community Collaboration (two items).

For each item, respondents are asked to select one of four response options, each corresponding to a readiness level from the equity matrix. These answer options are phrased using the exact language from the matrix to ensure alignment and interpretive consistency.

#### Scoring Logic and deployment

Each response is scored on a 1-to-4 scale, corresponding to the four readiness levels. All items are equally weighted. To enhance interpretability, the total readiness score is calculated as a percentage of the maximum possible score:

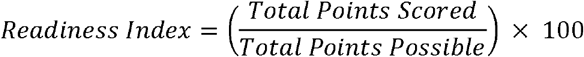

The resulting health equity readiness index is interpreted using the following thresholds:

- **25**–**37:** *Unrecognized Need*
- **38-62**: *Emerging Awareness*
- **53**–**87:** *Developing Capacity*
- **88**–**100:** *Established Practices*

In addition to an overall readiness score, dimension-specific scores are generated, enabling organizations to identify specific strengths and gaps. These results are visualized through a radar chart, allowing for intuitive interpretation and comparison across dimensions. Based on the results, a curated set of recommendations is provided to outline and close the delta to the next readiness level (see SM.3 for details).

The self-assessment tool is deployed as a web-based application [https://equityreadinessindex.replit.app/] that can be independently completed by DHOs. It is designed as a diagnostic entry point for identifying capacity gaps, supporting internal reflection, and informing subsequent planning, communication, or resource allocation efforts.

#### Tool application

To evaluate the health equity readiness self-assessment, we conducted a staged validation process combining qualitative feedback and structured survey data collection.

In the first stage, we conducted informal user testing with a convenience sample with our personal networks. Participants were asked to complete the assessment and provide open-ended feedback on language clarity, interpretability of the response options, and ease of use.

In the second stage, we conducted structured testing via Prolific (https://www.prolific.com/), a panel provider commonly applied in research studies [55]. In phase one, 30 participants were recruited to complete the self-assessment and provide feedback on the assessment tool through a follow-up survey. Their feedback informed minor grammatical corrections to the assessment questions. In phase two, an additional sample of 100 participants completed the assessment and a follow-up survey. We leveraged several pre-screening options from Prolific, namely age 18+, fluent English proficiency, no literacy difficulties (e.g., dyslexia), currently part- or full-time employment, medicine or information technology employment sector and health care and social assistance, information science and data processing, and medical/healthcare as industries. Additionally, we included a screening question at the beginning of the assessment tool to ensure current employment at a DHO. The follow-up survey consisted of four categories: (1) *demographic data* (age, gender, role, tenure, seniority, and self-reported health equity literacy), (2) *organizational characteristics* (operating country, number of employees, and type of digital health service offered), (3) *assessment evaluation* (items adapted from the IS success model [56], the Technology Acceptance Model (TAM) [57], and the Net Promoter Score; see SM.4 for details), and finally (4) *open-text feedback* to capture additional insights and suggestions for improvement. Combining the IS success model and the TAM is an established practice [58–61]. While TAM explains user beliefs and adoption, IS Success model contributes the quality antecedents and outcomes, yielding a coherent causal chain with strong explanatory power and content validity in tool evaluations [62]. Survey data was extracted, descriptive analysis was performed for the quantitative survey questions, and the free-text feedback underwent thematic analysis to identify overarching themes.

### Ethics

The study was exempt from a formal review and approval by the Ethics Committee of the University of St. Gallen in August 2025. Informed consent was obtained from all panelists before assessment.

## RESULTS

To address our research question, we developed a ***Health Equity Readiness Index***, a novel self-assessment tool for DHOs to assess their current readiness to implement health equity recommendations.

### Health Equity Readiness Matrix

The final readiness matrix (Table 1) is the foundation of this study, synthesizing conceptual frameworks from health equity, responsible AI, and digital innovation. It is composed of five dimensions that collectively define the organizational capabilities required to embed equity into the design, delivery, and governance of digital health interventions. Together, they provide a comprehensive framework to guide assessment and capacity-building.

**Table 1.**
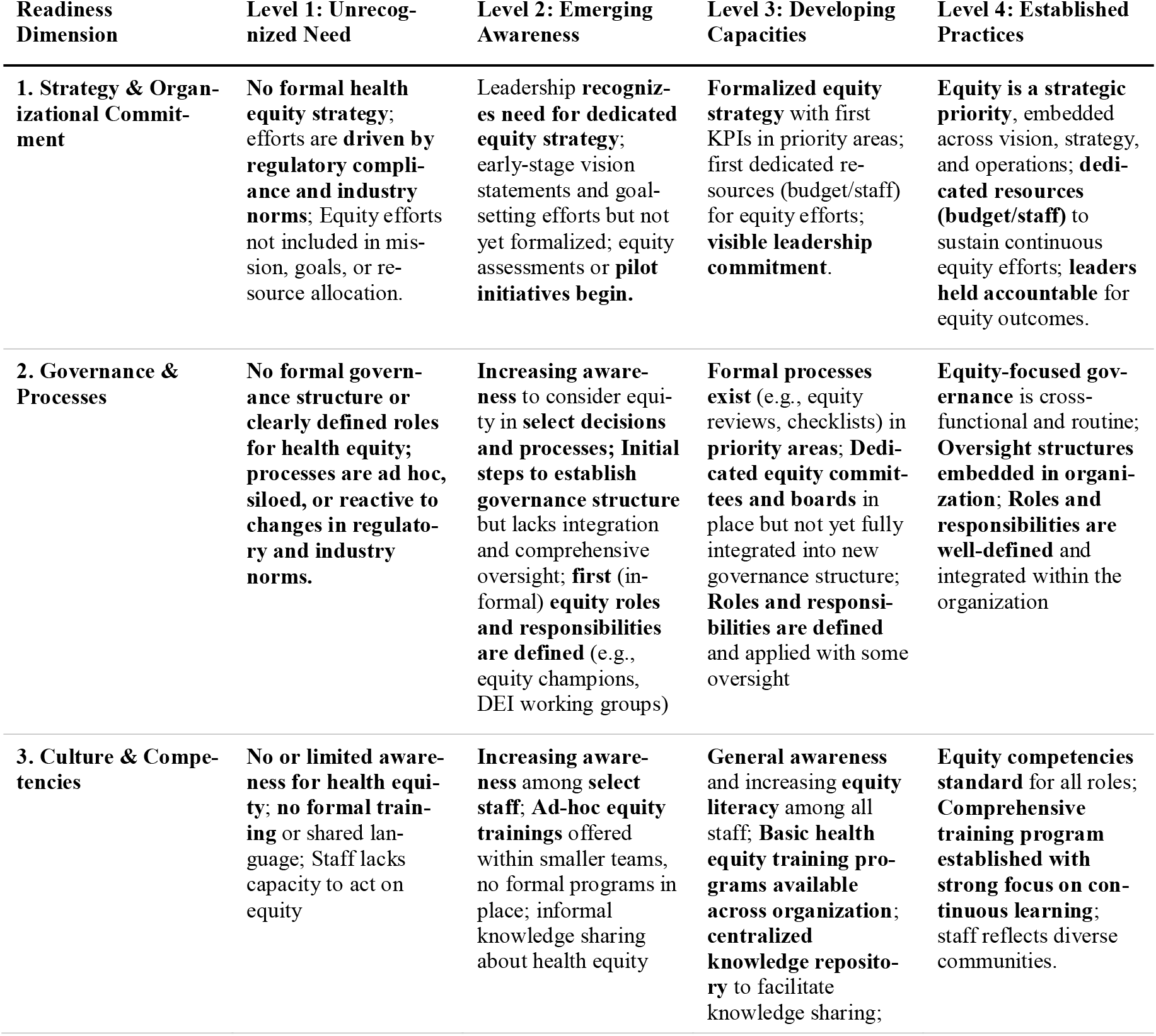
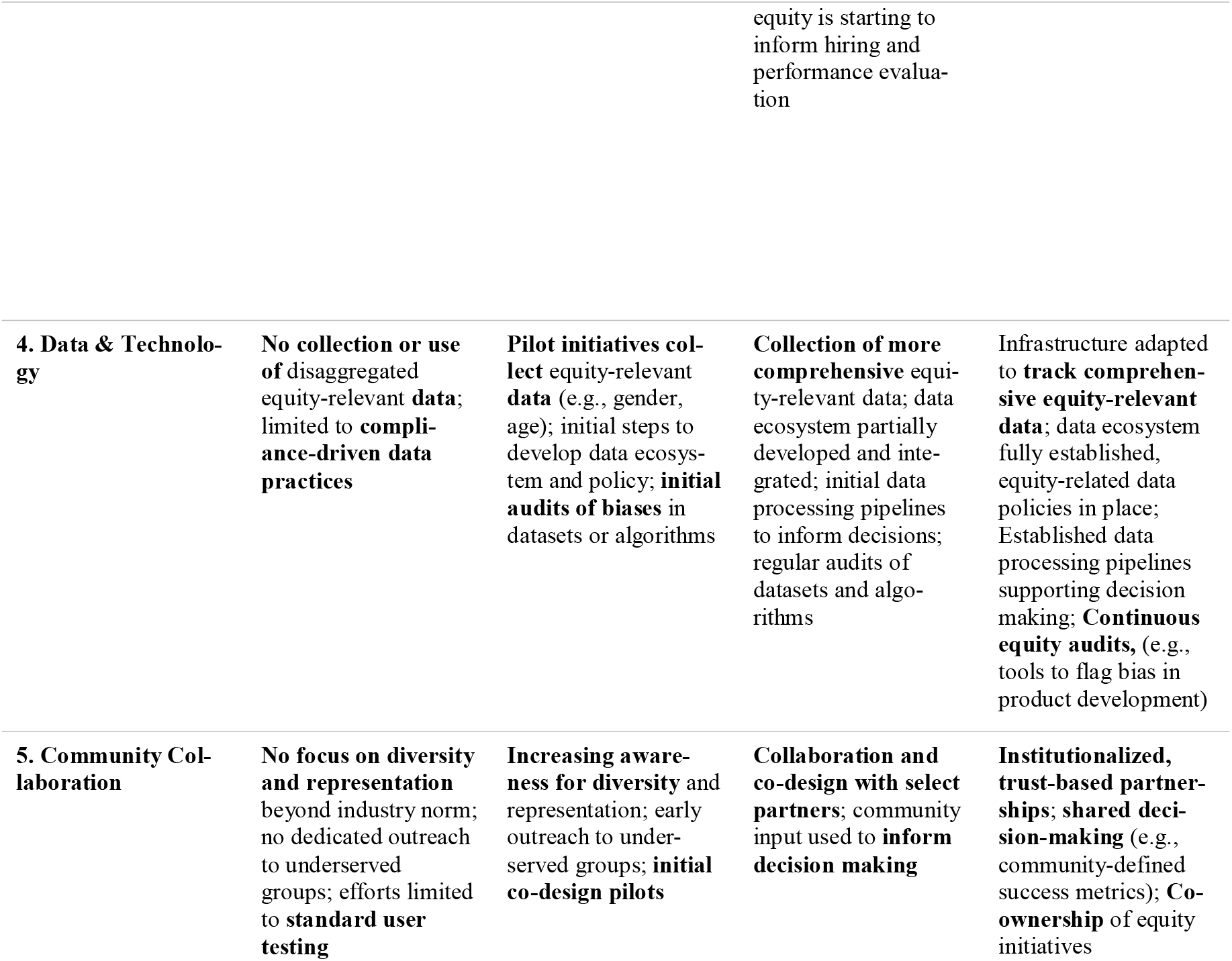
Health Equity Readiness Matrix: Populated matrix spanning five dimensions and four progressive levels of readiness. Each cell offers a descriptor of what readiness means for the respective level and dimension

#### Readiness Dimensions

The *Strategy & Organizational commitment* dimension focuses on the degree to which health equity is embedded into the strategic priorities and leadership structures of an organization. It encompasses the articulation of an explicit equity vision, the inclusion of equity in mission statements and goals, and the allocation of leadership attention and financial resources to equity initiatives. Organizations with high maturity in this dimension demonstrate clear accountability structures for equity, integrate equity into strategic planning cycles, and align resource deployment accordingly. This dimension provides the “why” that anchors all equity-related activities, ensuring they are not peripheral but integral to institutional purpose and direction.

The *Governance & Processes* dimension addresses the formal and informal mechanisms through which equity-related decisions are made and operationalized within an organization. It includes policies, protocols, review structures, and accountability systems that ensure equity considerations are consistently embedded into day-to-day operations and decision-making processes. High-functioning organizations in this domain establish cross-functional governance bodies for equity, routinely apply equity impact assessments, and incorporate equity metrics into performance management systems. This dimension serves as the operational backbone of equity integration, translating values into repeatable and auditable actions.

The *Culture & Competencies* dimension captures the internal values, norms, knowledge, and skills that shape how equity is understood and acted upon across the organization. It encompasses the degree to which staff at all levels are equipped to engage with equity, the presence of ongoing training programs, and the representation of diverse perspectives within teams. Organizations with higher maturity levels foster inclusive cultures, actively address bias, and invest in capacity-building initiatives that extend beyond compliance. Culture and competencies reflect an organization’s ability to transform intention into action by empowering its workforce to engage meaningfully with equity challenges.

The *Data & Technology* dimension refers to the infrastructure, tools, and analytical capabilities that enable organizations to collect, interpret, and act on equity-relevant information. It includes practices around data disaggregation (e.g., by race, ethnicity, gender, or geography), the use of equity-focused metrics, and the capacity to identify and mitigate algorithmic bias in digital solutions. This dimension bridges insight and action, enabling continuous monitoring and responsive adaptation to equity goals.

Finally, the *Community Collaboration* dimension assesses the extent to which organizations engage meaningfully with communities, particularly those historically underserved or marginalized. It includes mechanisms for incorporating community voices into strategy, co-design, evaluation, and governance processes. Higher levels of maturity are characterized by long-term, reciprocal relationships with trusted partners and structures that support shared decision-making and accountability. It ensures that equity initiatives are contextually relevant, socially legitimate, and responsive to real-world needs.

#### Readiness Levels

The matrix is further structured across four readiness levels that represent the evolving nature of organizational commitment and capability towards health equity, ranging from minimal recognition of equity concerns to full integration of equity across institutional functions.

The *Unrecognized Needs* level represents the most basic stage of readiness. At this level, health equity is not formally acknowledged as an organizational priority beyond compliance with external regulations, ethical codes, or industry norms. Organizations lack formal strategy, dedicated resources, and equity-relevant data practices. Efforts are often ad hoc or symbolic, and internal capacities for addressing equity are minimal or absent. Community engagement is minimal, non-strategic, or one-directional.

At the *Emerging Awareness* level, organizations begin to acknowledge health equity as an area of strategic importance with early leadership buy-in. While efforts may still be fragmented and limited in scope, pilot programs or one-time assessments are initiated to explore equity challenges within the organization or its services. First equity-relevant data collection begins in pilot initiatives and collaboration with community stakeholders is still exploratory, though growing interest in listening to underrepresented voices begins to shape early efforts.

The *Developing Capacity* level marks a turning point toward institutional change. Organizations at this stage take concrete steps to embed health equity into strategic planning, operations, and quality improvement efforts in priority areas. First dedicated roles, budgets, or working groups are introduced to advance equity goals. Equity-related data is collected more systematically and used to inform decision-making. Staff training on equity-related topics becomes more routine, and community engagement moves from consultation to co-creation in selected areas. While gaps remain, equity is no longer treated as an add-on but as a cross-cutting concern integrated into dedicated organizational learning and change processes.

The *Established Practices* level reflects organization-wide integration of health equity. Equity is embedded across all major functions, including strategy, governance, culture, data systems, and partnerships. The organization demonstrates sustained leadership commitment, continuous resource investment, and clear accountability structures. Equity metrics are incorporated into performance evaluation and decision-making at all levels. Staff are supported through ongoing training and competency development and community partnerships are reciprocal and institutionalized. At this stage, equity is not only a core value but a strategic advantage, reflected in the organization’s identity, operations, and impact.

### Assessment of the Digital Self-Assessment Tool

#### Respondent demographics

A total of 130 individuals completed the self-assessment survey, resulting in 124 valid data points (see Table 2 for full details). Respondents took an average of 21:21mins (median 18:34mins) to complete the full self-assessment, from instructions to filling out the feedback survey. They represented a range of age groups, with a mean age of 32.7 years (ranging 18–63). The sample included 56% female, 43% male, and 1% non-binary. Ethnicity data, provided by the sample provider, indicated that 26% of respondents identified as White, while 74% identified as Asian (6%), Black (61%), or Mixed (6%). This distribution suggests comparatively high representation of groups that are frequently underrepresented in health research.

**Table 2.** Respondent demographics of self-assessment survey.

| Characteristics | Number (n = 124) | Percentage |
| --- | --- | --- |
| **Respondent characteristics** |  |  |
| **Age** |  |  |
| 18 - 24 | 27 | 22% |
| 25 - 34 | 54 | 44% |
| 35 - 44 | 25 | 20% |
| 45 - 54 | 12 | 10% |
| 55+ | 6 | 5% |
| Average | 32.7 |  |
| **Gender** |  |  |
| Female | 70 | 56% |
| Male | 53 | 43% |
| Non-binary | 1 | 1% |
| **Ethnicity (simplified)** |  |  |
| White | 32 | 26% |
| Asian | 8 | 6% |
| Black | 76 | 61% |
| Mixed | 8 | 6% |
| **Seniority level** |  |  |
| Individual contributor | 32 | 26% |
| Middle management | 73 | 59% |
| Executive leadership | 19 | 15% |
| **Organizational tenure (in years)** |  |  |
| <1 | 4 | 3% |
| 1 - 3 | 58 | 47% |
| 4 - 6 | 37 | 30% |
| 7+ | 25 | 20% |
| **Equity literacy (self-reported)** |  |  |
| Very limited | 2 | 2% |
| Basic | 9 | 7% |
| Moderate | 38 | 31% |
| Strong | 63 | 51% |
| Expert | 12 | 10% |
| **Organizational characteristics** |  |  |
| **Number of employees** |  |  |
| <10 | 1 | 1% |
| 10 - 49 | 15 | 12% |
| 50 - 199 | 37 | 30% |
| 200 - 499 | 26 | 21% |
| 500 - 1000 | 18 | 15% |
| 1000+ | 27 | 22% |
| **Operating region** |  |  |
| Africa | 63 | 51% |
| America (North & South) | 13 | 10% |
| Asia | 3 | 2% |
| Europe | 40 | 32% |
| Global | 5 | 4% |
| **Digital Health Service Offering** |  |  |
| Electronic Health Record / Health IT Solutions | 24 | 19% |
| Health Data and Analytics Platforms | 22 | 18% |
| Health Equity or Social Determinants Solutions | 17 | 14% |
| Digital Therapeutics or Health Apps | 17 | 14% |
| Remote Care / Telemedicine Platforms | 16 | 13% |
| Medical Devices or Software as a Medical Device (SaMD) | 11 | 9% |
| Patient Engagement & Education Platforms | 11 | 9% |
| Clinical Decision Support Tools | 6 | 5% |

In terms of organizational role, respondents were primarily middle management (59%), followed by individual contributors (26%) and executive leadership (15%). Most had been with their organization for one to three years or longer (97%). Self-reported health equity literacy varied from very limited (2%) to expert (10%), with 91% reporting at least moderate literacy, likely reflecting both the prescreening for active digital health employment and the relevance of equity in this domain.

Respondents’ organizations also reflected diversity in size and scope. 13% were small organizations (<50 employees), 51% medium (50-499), and 36% large (>500). Geographically, organizations operated predominantly in Africa (51%), followed by Europe (32%) and the Americas (10%), while 4% reported global operations. Service offerings covered the breadth of the digital health ecosystem, with the most common being electronic health records/health IT solutions (n=24, 19%) and health data and analytics platforms (n=22, 18%). Other areas included health equity or social determinants solutions (14%), digital therapeutics or apps (14%), remote care/telemedicine platforms (13%), medical devices/Software as a Medical Device (9%), patient engagement platforms (9%), and clinical decision support tools (5%).

#### Perception of the self-assessment tool

Respondents rated the tool across six constructs: information quality, system quality, perceived usefulness, user satisfaction, net benefits, and advocacy (represented by the Net Promoter Score). Ratings were consistently high, with means ranging from 4.39 to 4.57 on a 5-point Likert scale ranging from strongly disagree (1) to strongly agree (5) (Table 3). Advocacy received the highest score (M=4.57, SD=0.67), followed closely by information quality (M=4.56, SD=0.61). Even the lowest-rated construct, perceived usefulness, remained well above 4.3 (M=4.39, SD=0.55). Internal consistency was strong across multi-item constructs (Cronbach’s α = 0.75–0.87). These findings suggest that respondents not only found the tool easy to use but also saw value in its capacity to inform organizational reflection and action.

**Table 3.** Construct overview.

| Construct | # Items | Cronbach's $\alpha$ | Mean (M) | Standard deviation (SD) | Adapted from |
| --- | --- | --- | --- | --- | --- |
| Information quality | 2 | 0.87 | 4.56 | 0.61 | IS Success Model |
| System quality | 2 | 0.75 | 4.55 | 0.58 | IS Success Model |
| Perceived usefulness | 3 | 0.87 | 4.39 | 0.55 | TAM |
| User satisfaction | 2 | 0.85 | 4.49 | 0.64 | IS Success Model, TAM |
| Net benefits | 2 | 0.77 | 4.42 | 0.63 | IS Success Model |
| Advocacy | 1 | - | 4.57 | 0.67 | Net promoter Score |

In addition to the quantitative feedback, respondents provided rich free-text feedback on what they found most useful and where the tool could be improved. Thematic analysis yielded eight recurring themes, grouped into strengths (five themes) and improvement suggestions (three themes). Table 4 presents an overview of the themes with descriptions, number of mentions, and illustrative quotes.

**Table 4.**
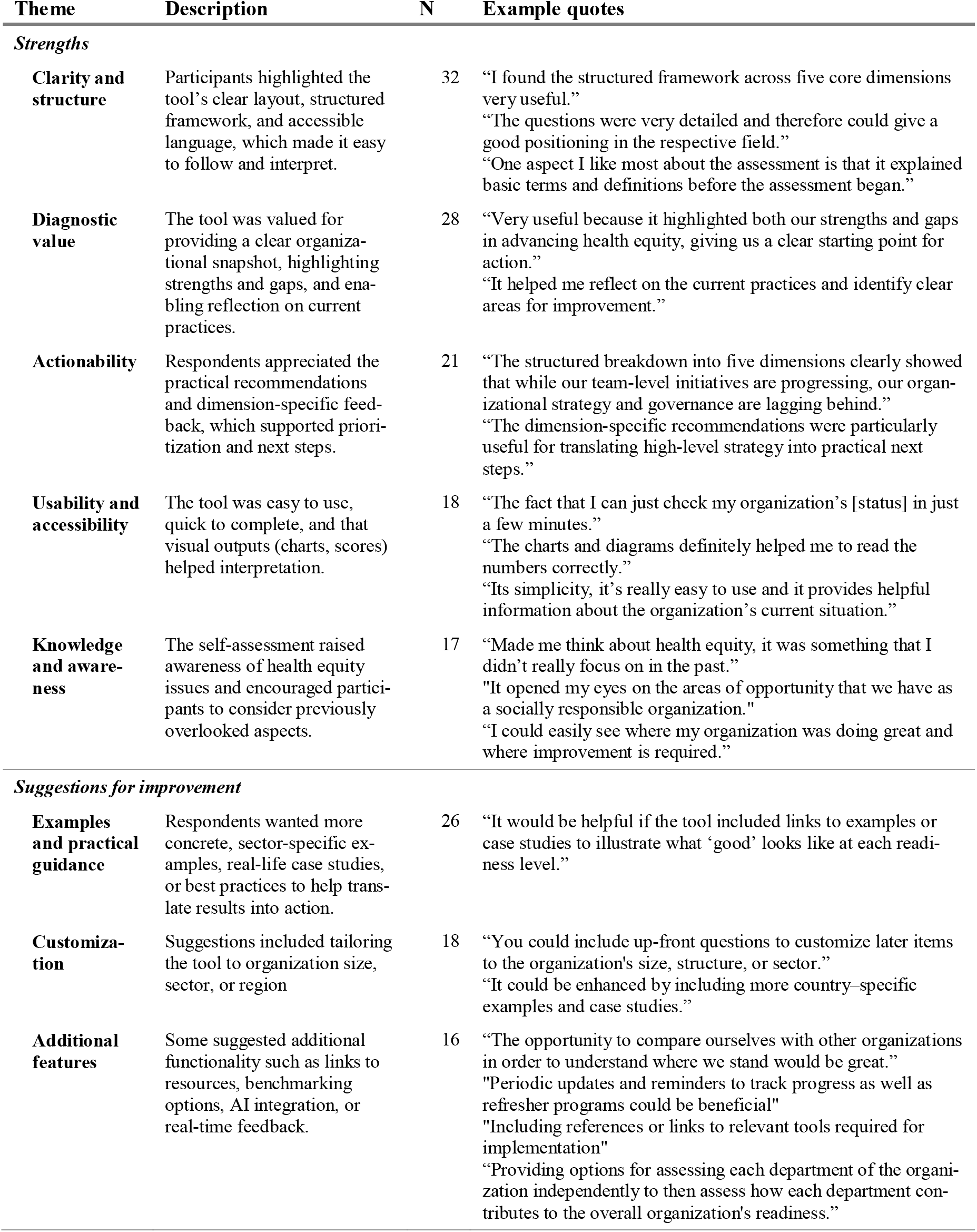
Synthesis of qualitative feedback: Results of thematic analysis grouped in strengths and suggestions for improvement

Participants most frequently emphasized the tool’s *clarity and structure*, noting that its logical flow and clear definitions made it straightforward to complete. It was also praised for its *diagnostic value*, with many reporting that it helped them identify organizational blind spots and gain a holistic overview of readiness. Beyond diagnosis, respondents valued the tool’s *actionability*, especially the dimension-specific recommendations that supported concrete next steps rather than merely producing a static score. Its *usability and accessibility* were also highlighted, with respondents finding it quick to complete, visually intuitive, and relevant across roles. Finally, some emphasized that it fostered *knowledge and awareness*, prompting reflection on equity considerations that might otherwise be overlooked.

At the same time, respondents identified opportunities for further enhancement. The most common request was for more *examples and practical guidance*, including case studies and sector-specific illustrations of “what good looks like” at different readiness levels. Others suggested greater *customization*, such as tailoring content to organizational size, sector, or region, and offering benchmarking or progress-tracking features. Finally, suggestions focused on *additional features*, including links to external resources, benchmarking options, and optional department-level assessments. Notably, a considerable number of respondents (n=30) stated explicitly that no improvements were needed, describing the tool as already intuitive and effective.

Overall, the feedback suggests that the self-assessment tool effectively combines structured orientation with reflective prompts that help organizations critically assess their readiness. Its recommendations and feedback features were perceived as particularly valuable in translating assessment results into actionable steps. While respondents appreciated its clarity and accessibility, they also identified opportunities to strengthen the tool’s practical impact through greater customization, illustrative case examples, and additional features. Importantly, a large proportion of participants expressed high satisfaction, reinforcing the tool’s perceived relevance and usability in digital health contexts.

## DISCUSSION

This study sought to advance the operationalization of health equity within DHOs by developing and testing a practical readiness assessment tool. Drawing on established readiness model frameworks on health equity, organizational change, responsible AI, and digital innovation literature, we created the *Health Equity Readiness Index* and a corresponding self-assessment tool. The 15-items tool enables DHOs to evaluate their current level of equity readiness across five key dimensions. The tool was evaluated in a sample of 124 professionals working across diverse digital health contexts, combining quantitative ratings with qualitative feedback on usability, interpretability, and relevance.

### Theoretical contributions

Our findings extend readiness model scholarship into the underexplored domain of health equity. While readiness models are well-established in fields like digital transformation [41], cybersecurity [42], and (responsible) AI [43, 44], equivalent approaches remain limited for health equity, particularly within digital health. Our study fills this gap by proposing a structured, theory-informed framework specifically tailored to DHOs. The *Health Equity Readiness Index* defines health equity readiness not as a binary state but as a developmental trajectory from regulatory compliance to equity leadership, offering conceptual clarity on what “readiness” for health equity entails. It contributes a new lens to the discourse on organizational capacity, bridging knowledge from responsible innovation and equity frameworks. By embedding health equity into DHOs, the framework helps to move beyond rhetorical commitments and supports the measurable integration of equity into decision-making and operations. It further helps define what constitutes meaningful progress in a domain where measurement is often elusive or contested.

This framework also contributes to the emerging scholarship on equity-centered innovation by offering an organizational lens through which readiness can be diagnosed, tracked, and strengthened over time. It complements the “equity by design” paradigm [27] by situating design within broader organizational systems, emphasizing that equitable outcomes depend not only on product features but also on the readiness of the institutions creating and deploying them. The positive reception of the self-assessment suggests that embedding health equity into strategy, governance, culture, data, and community relations is not only theoretically compelling but also practically meaningful to DHOs.

### Practical implications

For DHOs, the self-assessment tool serves multiple practical functions. It offers a low-barrier entry point for initiating and structuring equity discussions. Respondents emphasized its clarity and diagnostic capacity, suggesting its potential to reveal blind spots and support prioritization of equity initiatives. Dimension-specific scores and recommendations may help organizations allocate resources strategically, align with funder and regulator expectations, and use results for internal advocacy.

Thereby, the tool complements established assessment approaches in digital health. Widely used frameworks such as the HIMSS Digital Health Indicator [63] or the NHS Digital Technology Assessment Criteria (DTAC) [64] evaluate infrastructure, interoperability, safety, and usability. However, they do not assess whether organizations are systematically prepared to produce equitable outcomes. The strong usability and relevance ratings in our survey suggest that integrating equity as a core dimension of organizational maturity is both feasible and valued in practice.

At the same time, our work extends equity-focused assessment tools that operate at other levels of the health system. For example, the WHO Health Equity Assessment Toolkit (HEAT) [65] enables stakeholders to explore and report population-level health inequalities, and national health literacy surveys, such as the German HLS-GER [8], demonstrate persistent disparities in individuals’ ability to understand and use health information. While these instruments help characterize the problem space, they do not evaluate the organizational conditions that shape equitable digital innovation. By shifting the analytical lens from population capability to institutional capacity, the *Health Equity Readiness Index* fills a meso-level gap between public health objectives and the organizational systems responsible for operationalizing them. Given the dual responsibility of DHOs, as both health service providers and technology developers, they have an outsized influence on how health equity challenges are addressed in the digital age. The readiness tool reflects this by encouraging organizations to see themselves not only as passive implementers of public health goals but as active agents shaping equity outcomes through strategy, governance, culture, data, and community relationships.

Beyond individual organizations, the tool holds potential value for funders, policymakers, and health system actors. By operationalizing equity readiness into measurable levels, it could inform eligibility criteria for grants, certification programs, or reimbursement schemes. For example, funders could require a minimum readiness level for participation in innovation programs, or policymakers could link readiness assessments to reimbursement eligibility under public health systems. It may also support regional capacity-building by identifying structural inequities in digital health ecosystems. As such, the readiness framework may not only support organizational transformation but also contribute to the institutionalization of health equity.

### Limitations and future research

While promising, the approach presented in this study has several limitations that must be acknowledged. The tool was designed as a self-administered, individual-level diagnostic. While this design supports low-barrier uptake, it may also introduce individual bias or fail to capture the diversity of perspectives within a given organization. Future iterations may explore facilitated team-based assessments, integration into organizational learning processes, or use in workshop settings to foster cross-functional dialogue. Moreover, the tool was developed with a generalizable structure, but readiness for health equity may manifest differently across settings, health system contexts, and regulatory environments. For instance, small startups operating in low-resource settings may face different barriers than well-funded platforms in tightly regulated markets. As such, future research could investigate contextual adaptations for specific subdomains (e.g., mental health, chronic disease management, AI-based diagnostics) to increase relevance and precision.

Finally, it is worth noting that readiness is not a static or neutral concept. It reflects not only organizational capacity but also normative judgments about what constitutes legitimate progress. While the tool attempts to define levels in a developmental sequence, real-world progress is often non-linear and influenced by external shocks, shifting leadership priorities, or political pressures. Moreover, self-assessments may be subject to social desirability bias, overconfidence, or misalignment between perceived and actual capability. To mitigate this, future research may explore triangulation with third-party assessments, peer review, or integration into broader equity audits. While the readiness matrix and assessment tool were specifically developed with DHOs in mind, the underlying organizational dimensions and developmental logic may be transferable to other actors in the health innovation ecosystem, including healthcare providers, funders, research consortia, and public health institutions. Future studies could explore how well the framework applies beyond DHOs and what contextual adaptations may be necessary to increase its relevance and impact in other organizational settings. Such extensions could enhance the tool’s utility as a sector-wide standard for advancing organizational readiness for health equity.

Despite these limitations, the *Health Equity Readiness Index*, the underlying matrix and supporting self-assessment tool represent a foundational step toward embedding health equity more deeply into DHOs. By enabling organizations to benchmark, reflect, and act, the tool helps build the internal conditions necessary for equity to move from aspiration to implementation.

## Data Availability

All data produced in the present study are available upon reasonable request to the corresponding author

## CONTRIBUTIONS

*Author1* developed the research questions and conceptual design of this research with input from *Author 2* and *Author3. Author1* performed the analysis of relevant literature and proposed an initial version of the readiness matrix, which was finalized in a co-design workshop among all authors. *Author1* developed the self-assessment tool, performed the survey analysis, and initiated the manuscript drafting, incorporating valuable feedback from *Author2* and *Author3* throughout the iterations. The final version received full approval from all authors.

## DECLARATION OF GENERATIVE AI AND AI-ASSISTED TECHNOLOGIES IN THE WRITING PROCESS

During the preparation of this work the authors used ChatGPT in order to optimize language and grammar. After using the tool, the authors reviewed and edited the content as needed and take full responsibility for the content of the publication.

