## Supplementary Materials for "Equity Readiness Index for Digital Health Organizations"

*Supplementary Materials (SM)*

SM.1: Matrix population: sample descriptors

Sample descriptors for the dimension *Strategy & Organizational Commitment* to visualize matrix population.

|  | **Level 1** | **Level 2** | **Level 3** | **Level 4** |
| --- | --- | --- | --- | --- |
| **Reference Work** *Willems, 2025 [1]* | There is no vision or goal for Responsible AI. The organization continues to operate as usual without specific AI-related objectives. There is no roadmap. The organization is in the early stages of exploring AI and its potential applications. [...] AI initiatives are ad-hoc and lack oversight. | Initial awareness of the need for a Responsible AI vision and goals, but they are not yet formulated. The distinction between strategy and tactical is not clear. There is a growing recognition that AI can facilitate change. Initial discussions and plans are being made, but a structured roadmap is not yet in place. | AI vision and goals are defined but not yet fully communicated or integrated across the organization. [...] initial steps are taken to align AI initiatives with organizational goals. A preliminary AI roadmap exists, outlining key initiatives and milestones. The roadmap begins to address identified challenges and suggests how the transition to AI should be made responsibly. | Responsible AI vision and goals are well-defined and communicated, with alignment to broader organizational objectives. There is a process in place to update the vision and goals, considering changes in the organization and the external environment. A detailed AI roadmap is in place, [...] integrated with business strategy and is regularly updated. |
| **Health Equity Lens Adoption** | **No formal health equity strategy**; efforts are **driven by regulatory compliance and industry norms**; Equity efforts not included in mission, goals, or resource allocation. | Leadership **recognizes need for dedicated equity strategy**; early-stage vision statements and goal-setting efforts but not yet formalized; equity assessments or **pilot initiatives begin.** | **Formalized equity strategy** with first KPIs in priority areas; first dedicated resources (budget/staff) for equity efforts; **visible leadership commitment**. | **Equity is a strategic priority**, embedded across vision, strategy, and operations; **dedicated resources (budget/staff)** to sustain continuous equity efforts; **leaders held accountable** for equity outcomes. |

SM.2: Self-assessment items

|  |  | **Answer options** | | | |
| --- | --- | --- | --- | --- | --- |
| **Dimension** | **Question** | **Unrecognized** | **Emerging** | **Developing** | **Established** |
| Strategy & Organizational Commitment | To what extent is health equity integrated into the organization’s strategy, mission, and goals? | No formal strategy exists for health equity. It is not reflected in the organization’s mission, goals, or resource allocations. | Leadership recognizes the need for a dedicated equity strategy. Early-stage vision statements and goal-setting efforts exist but are not yet formalized. | A formal equity strategy is in place, with initial KPIs and efforts to operationalize equity in select strategic areas. | Equity is a clearly defined strategic priority, fully embedded across vision, strategy, and operations. |
|  | How committed and accountable is leadership for advancing health equity in your organization? | Leadership involvement in health equity is minimal, with no visible commitment or ownership. | Leadership has begun articulating a vision for equity, but no accountability mechanisms are in place. | Leadership demonstrates visible commitment to equity with emerging mechanisms for accountability. | Leaders are held accountable for equity outcomes, and their sustained commitment is reflected in decisions and actions. |
|  | What resources and actions has your organization dedicated to implementing health equity efforts? | No dedicated resources (budget or staff) are allocated to equity. Activities are limited to meeting regulatory requirements or industry standards. | Pilot equity initiatives or assessments are underway, but resources are limited, short-term, or inconsistently allocated. | First dedicated budget and staff are assigned to equity efforts, enabling more structured and ongoing implementation. | Sustained equity-focused resources (budget and staff) are embedded across the organization, supporting continuous efforts and measurable outcomes. |
| Governance & Processes | How well-developed is your organization’s governance structure for supporting health equity? | No formal or informal governance structure exists to oversee health equity efforts. | Initial steps to form a governance structure are underway, but efforts are limited, siloed, or lack comprehensive oversight | Equity-focused committees or boards exist and provide guidance, but are not yet fully embedded in organizational governance. | Equity governance is fully embedded, cross-functional, and routinely informs decision-making across the organization. |
|  | How clearly defined and integrated are roles and responsibilities for advancing health equity in your organization? | There are no defined roles or responsibilities for health equity, any efforts are informal or ad hoc. | Informal roles such as equity champions or DEI working groups exist, but responsibilities are not formalized or applied organization-wide. | Roles and responsibilities are formally defined, with initial oversight mechanisms and partial organizational integration. | Roles and responsibilities are formalized, consistently applied across the organization, and supported by clear accountability mechanisms. |
|  | How formalized and consistent are your organization’s processes for overseeing and ensuring health equity? | Processes related to health equity are ad hoc, siloed, or reactive to compliance needs; no formal oversight exists. | Initial steps are taken to establish oversight mechanisms, but they are limited in scope and not yet comprehensive; equity considerations are included in select processes. | Formal processes such as equity reviews or checklists exist in priority areas, and oversight is applied inconsistently but growing. | Oversight mechanisms and equity-focused processes are routine, organization-wide, and embedded in decision-making and accountability systems. |
| Culture & Capabilities | How widespread is awareness and understanding of health equity across your organization? | Awareness of health equity is minimal or limited to isolated individuals. | Awareness is growing among select staff. Equity is occasionally discussed in small teams or informal settings. | Awareness and equity literacy are increasing across the organization, with a shared understanding beginning to form. | Health equity awareness is embedded organization-wide amd staff demonstrate shared understanding and commitment to equity values in their work. |
|  | To what extent does your organization provide training and capacity-building opportunities to advance health equity? | No formal training programs or resources for health equity. Staff are not equipped to address equity-related issues. | Ad-hoc trainings are provided in some teams, but there is no structured or organization-wide program. | Basic training programs are available across the organization, and resources such as toolkits or repositories support knowledge sharing. | Comprehensive, ongoing training is embedded across roles and functions, emphasizing continuous learning and applied skill-building. |
|  | To what extend does your organization align staffing decisions and workforce diversity with health equity goals? | Equity is not considered in staffing decisions, and workforce diversity is not actively prioritized. | Initial efforts to consider equity in staffing are underway. Discussions about workforce diversity have begun but are not formalized. | Increasing efforts to include dedicated equity goals in staffing decisions and the workforce is becoming more reflective of the communities served. | Workforce diversity reflects the communities served, and equity is consistently embedded in recruitment, hiring, and retention practices. |
| Data & Technology | To what extent does your organization collect and disaggregate equity-relevant data? | Equity-relevant data is not collected or disaggregated. | Pilot initiatives collect limited equity-relevant data (e.g., gender, age) | Collection of more comprehensive equity-relevant data is underway and data is disaggregated and covers a broader range of equity dimensions. | Comprehensive, disaggregated equity data is routinely collected across relevant dimensions and used to generate actionable insights. |
|  | How developed is your organization’s infrastructure for managing, storing, and accessing equity-relevant data? | No data ecosystem or infrastructure in place; data practices are limited and compliance-driven. | Initial steps to develop a data ecosystem and policy are underway | A partially developed and integrated data ecosystem is in place, supporting data collection, management, and use | A fully established and integrated data ecosystem supports equity-relevant data collection, management, and use. Robust policies and infrastructure are in place to ensure data quality and accessibility. |
|  | To what extent does your organization evaluate datasets and algorithms for equity-related bias? | Bias in data or algorithms is not evaluated beyond what is legally required. | Initial evaluations of bias in datasets or algorithms are conducted in limited areas or using basic methods. | Bias evaluations are conducted regularly using defined methods, with results informing mitigation strategies. | Continuous and systematic audits are conducted using advanced tools, with results actively used to mitigate bias in both data and product development. |
|  | How consistently does your organization use equity-relevant data to inform strategic and operational decisions? | Equity-relevant data is not used to guide decisions or evaluate outcomes. | Equity-relevant data informs decision-making in select pilot projects, but use is not yet widespread. | Partially developed data pipelines support equity-relevant decision-making in some departments or processes. | Robust data pipelines routinely support decision-making across the organization, enabling tracking and achievement of equity-related goals. |
| Community Collaboration | How actively does your organization engage with marginalized communities to build long-term relationships? | No dedicated outreach or engagement with marginalized communities. | Initial outreach to marginalized communities is underway, but relationships are limited and not sustained. | Initial partnerships with select marginalized communities have been established, with some efforts to build trust and reciprocity. | Trust-based, long-term partnerships with marginalized communities are in place. |
|  | To what extent does your organization collaborate and co-design with communities and partners to to shape priorities and solutions? | No collaboration or co-design with communities beyond conventional user testing or surveys. | Initial co-design activities are being piloted with limited, consultative community involvement. | Ongoing co-design efforts with select community partners, with input informing program design and implementation. | Collaboration and co-design are deeply embedded, with shared decision-making, community-defined success metrics, and co-ownership of equity initiatives. |

SM.3: Self-assessment tool

Below you can find an exemplary overview of self-assessment visualization, organized along the flow through the tool. This includes the introduction page, outlining the motivation for and the structure of the index. This is then followed by assessment questions, represented here by the first question of each dimension to showcase the progression. Finally, this concludes at the results page, showcasing the overall score, the spiderweb visualization and the supporting information and recommendations. The full tool here: <https://equityreadinessindex.replit.app>

SM.3.1: Introduction page

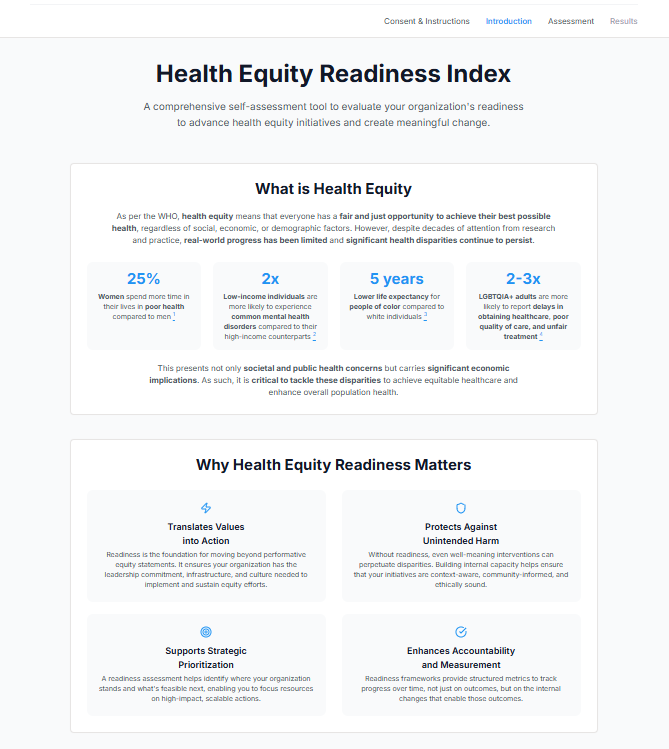

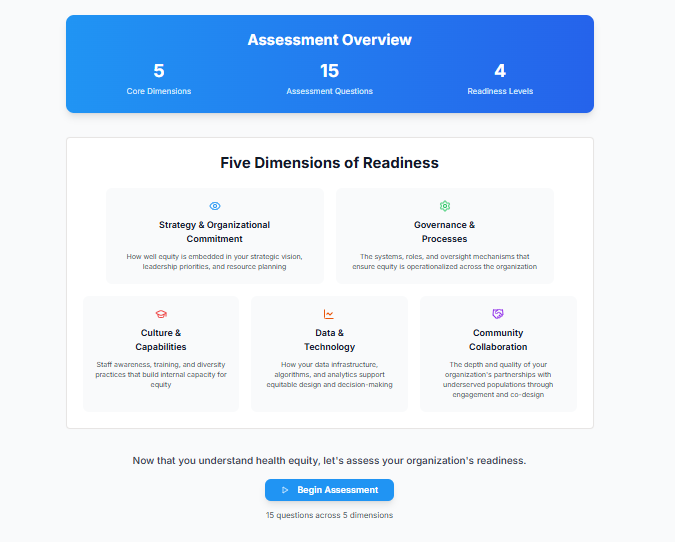

SM3.2: Self-assessment items (selective examples per dimension)

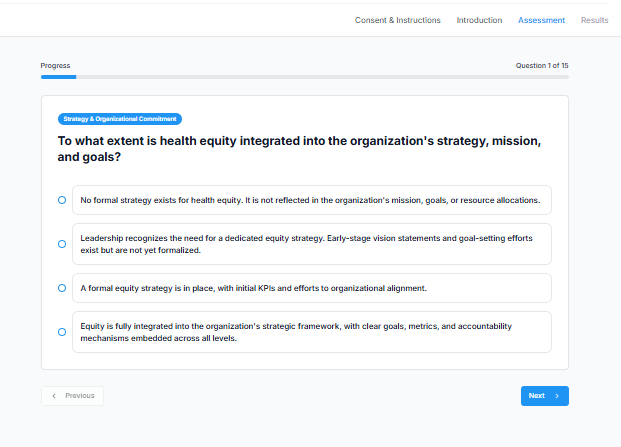

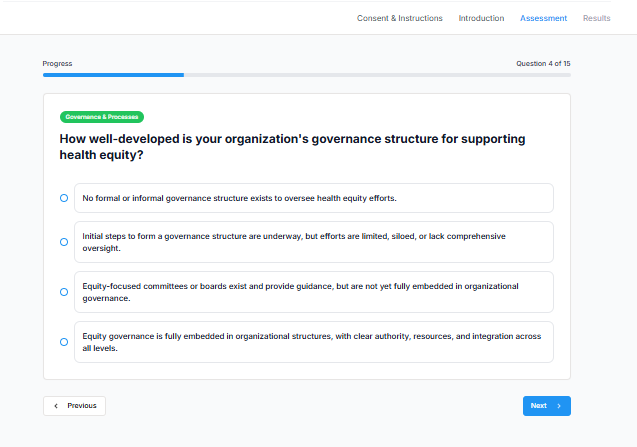

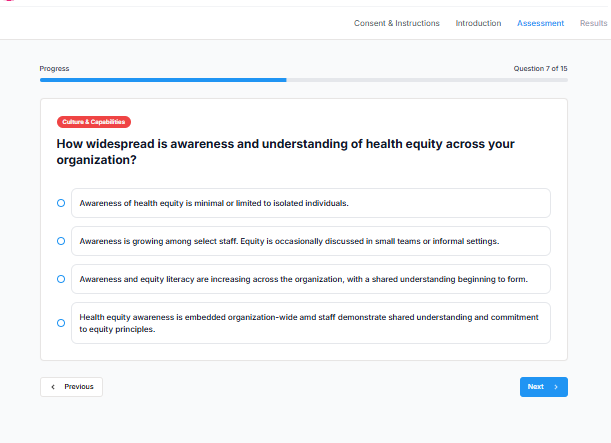

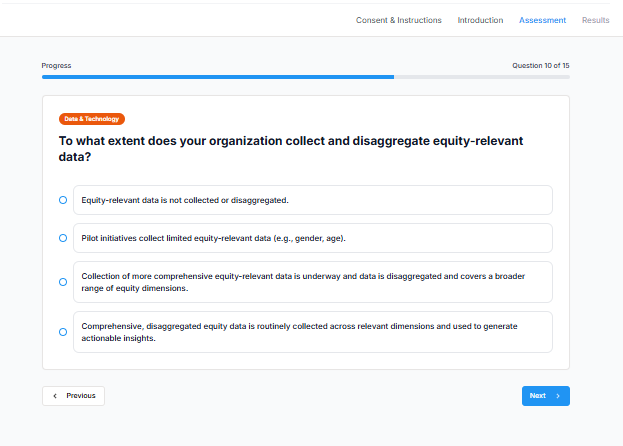

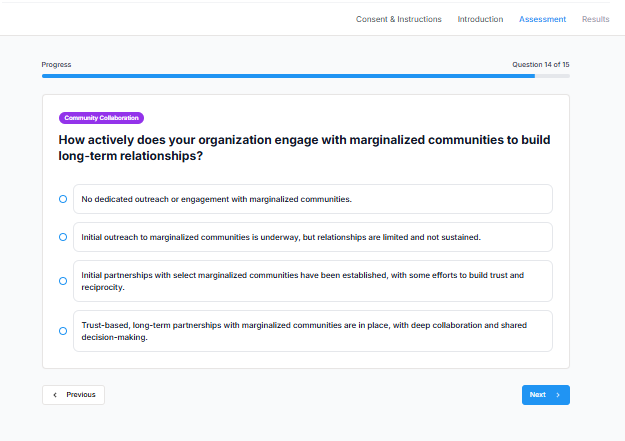

SM3.3: Results page

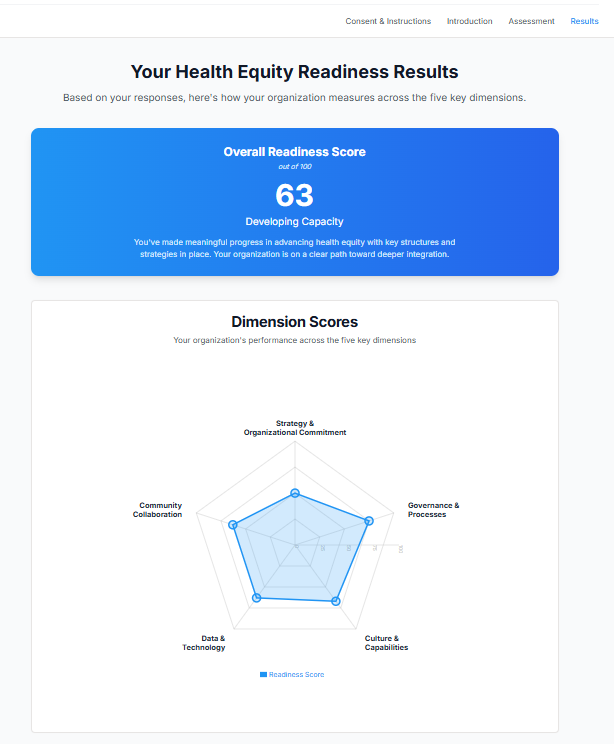

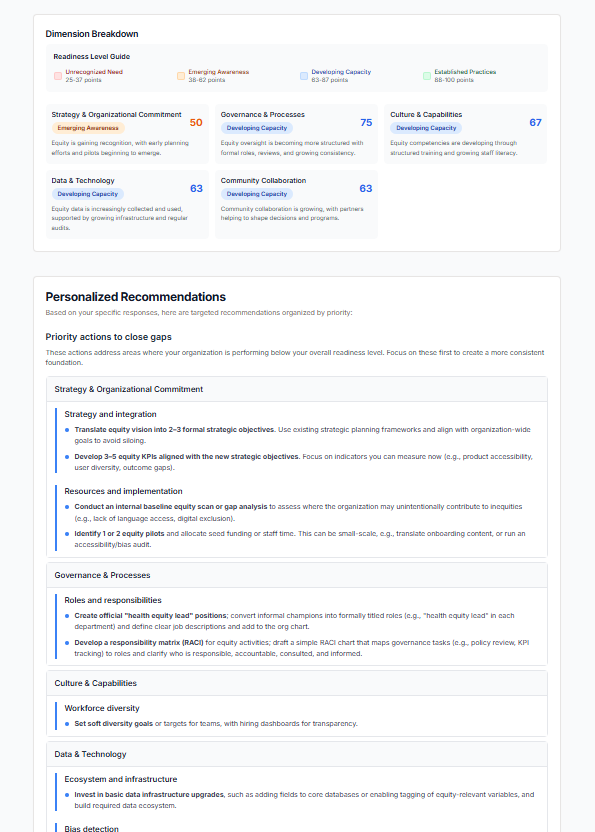

SM.4: Evaluation of the self-assessment tool

| **Construct** | **Question** | **Adapted from** |
| --- | --- | --- |
| Information Quality | The questions and answer options were clear and easy to understand | IS Success Model [2] |
|  | The language used was appropriate for my level of expertise. |  |
| System Quality | The length of the assessment was appropriate. | IS Success Model [2] |
|  | The tool was easy and intuitive to use. |  |
| Perceived Usefulness | The self-assessment reflected challenges and priorities relevant to my organization. | TAM [3] |
|  | The dimension-specific results and recommendations offered helpful insight. |  |
|  | The recommendations provided were actionable and realistic. |  |
| User Satisfaction | I found the self-assessment valuable for our organization. | IS Success Model [2], TAM [3] |
|  | I would consider using this tool again in 6–12 months to track progress. |  |
| Net Benefits | I feel better equipped to take the next steps on our equity journey. | IS Success Model [2] |
|  | The assessment helped me better understand where my organization stands on health equity readiness |  |
| Advocacy | I would recommend this tool to others in digital health organizations. | Net Promoter Score (NPS) |
